# Timing of antenatal hemoglobin testing and associated maternal and neonatal characteristics at public hospitals in Lusaka, Zambia

**DOI:** 10.1101/2025.02.11.25322071

**Authors:** Adenike Oluwakemi Ogah, Chrispin Mwando, Kenneth Chanda, Selia Nganjo

## Abstract

**Background:** The identification and mitigation of anaemia during pregnancy constitutes a significant public health priority. The assessment of hemoglobin values during antenatal is intended to be a standard practice. However, this may not reflect the actual practices observed in public hospitals within developing countries.

**Subject and methods:** This is a secondary analysis of a cross-sectional study of 489 mother-singleton, term newborn pairs, who were consecutively recruited from the admission wards of six public hospitals in Lusaka, Zambia. The presence or absence and timings of antenatal hemoglobin testing documented in the maternal hospital records was utilized as a metric to evaluate the quality of antenatal care provided. The relationship between the variables was explored using Chi-square tests and a binary logistic regression model. The findings were reported in terms of p-values, odds ratios, and 95% confidence intervals.

**Results:** Maternal antenatal hemoglobin data were obtained from the hospital records of 239 (48.9%) out of the 489 mothers in the study. Of these 239, 156 (65.3%) maternal most recent hemoglobin levels were determined more than 4 weeks before delivery and only 83 (34.7%) were determined within 4wks prior to delivery. Additionally, 250 (51.1%) of the 489 mothers had no file record of antenatal hemoglobin testing. An increase in maternal body mass index was associated with a decreased likelihood of missing antenatal hemoglobin records (p<0.001; OR=0.468; [95%CI 0.355, 0.617]).

Mothers with a documented history of previous miscarriage, low antenatal hemoglobin levels (Hb < 11 g/dL), underlying medical conditions, lower level of education and babies with low hemoglobin (<15g/dl) were more likely to have their most recent antenatal hemoglobin test conducted more than four weeks prior to delivery; OR 2.65 (p=0.015; 95% CI 1.211, 5.780); OR 2.22 (p=0.010; 95% CI 1.213, 4.070); OR 4.612 (95%CI 1.342, 15.851); OR 2.182 (95%CI 0.593, 8.022) and (p<0.001; OR 2.89; 95%CI 1.590, 5.236), respectively. The analysis revealed that mothers with more than one prior miscarriage (18 out of 26, accounting for 69.2%, p = 0.018) were 3.134 times more likely (95% CI: 1.195, 8.218) to belong to the group of mothers lacking documented antenatal hemoglobin test.

HIV rate in pregnancy was 3.9% in this study. All (except diabetes mellitus) the medical conditions documented were either associated with remote (more than 4weeks) or no hemoglobin testing before delivery.

**Conclusion:** The significant proportion of mothers who did not undergo antenatal hemoglobin testing, coupled with those whose hemoglobin levels were assessed remotely, indicates a deficiency in antenatal care within the study population. Mothers, who presented with absent or remotely conducted antenatal hemoglobin assessments were more likely to exhibit characteristics such as low body mass index (BMI), reduced hemoglobin levels, a history of previous miscarriages, lower educational attainment, and underlying medical conditions, as well as giving birth to newborns with low hemoglobin levels. Consequently, it is imperative that these specific groups of mothers receive closer monitoring. There exists an urgent need to enhance the standard of antenatal care provided in public health facilities to prevent adverse maternal and neonatal health outcomes.

## Background

Anemia is common in pregnancy, occurring in almost 40% of pregnant mothers globally, while newborn anemia in Sub-Saharan Africa is between 25 and 30%.^1^ Anemia increases in frequency with each trimester, according Zmei et al., USA^2^ Antenatal anemia is therefore associated with higher odds of several adverse maternal and neonatal outcomes, such as transfusion, cesarean delivery, and maternal death,^3,4,5^ and poor neonatal outcomes, including preterm birth, low birthweight, small-for-gestational-age infants, and neonatal and perinatal death^6,7,8,9^ Postpartum depression, inadequate maternal-infant interaction, and suboptimal neonatal psychomotor scoring are significant anemia outcomes that also warrant attention.^6,7,8^ Tissue hypoxia, delayed brain development, stunted growth, chronic heart failure, infectious diseases such as HIV and hepatitis as a result of repeated blood product infusions, and eventually multiple organ failure are all complications of unrecognized and untreated anemia in infants.^9,10,11,12,13^

In a prospective cohort study of 352 pregnant mothers in South western Uganda, Ngozi et al. found a 17% prevalence of newborn anemia based on umbilical cord blood and hemoglobin levels <13g/dl.14 The Uganda study identified maternal anemia, cesarean delivery, high maternal parity, and young maternal age as risk factors for neonatal anemia. In Ethiopia, 22% of pregnant women were found to be anemic.^15,16^

Infants under six months of age are at significant risk of anemia due to their rapid growth and insufficient iron intake, as breast milk is poor in iron.^17^ Consequently, these infants primarily depend on iron derived from intrauterine life. The situation becomes considerably more concerning when the pregnant mother, who serves as the main source of fetal iron, experiences anemia. Therefore, it is imperative that all women, particularly in developing countries, receive routine hemoglobin testing, consistent dietary guidance as well as oral iron supplements throughout their pregnancy. This approach is essential to optimize iron intake and prevent anemia.^18^ There is insufficient data and therefore no consensus, to determine the most appropriate time for anemia screening in pregnancy and follow-up. Though the American College of Obstetricians and Gynecologists’ Committee (ACOG) endorses universal screening for anemia in pregnant mothers with a complete blood count in the first trimester and again in the early third trimester, this guideline provides no exact timing or schedule for screening.^19^ The National Institute for Health and Care Excellence (NICE) guidelines further advised hemoglobin testing at the beginning of pregnancy (or at booking) and then at 28 weeks gestation; however, this also lacks sufficient evidence to support this recommendation.^20,21,22^

Yefet et al. in a secondary analysis of a prospective cohort data, which assessed the efficacy of a routine screening protocol for postpartum anemia diagnosis, between 2015 and 2016, at the Emek Medical Center in Israel, demonstrated that Hb screening between 30 and 36 gestational weeks is a strong predictor for anemia at delivery, and therefore suggested pre-delivery Hb testing at 24–30 weeks and then four weeks later.^23^

Contrary to Zmei et al, Sherard et al., North Carolina, USA (2001) in a prospective study, observed a progressive decline in the frequency of anemia from 20% at 26-28 weeks to 11% at term; and argued that if the value obtained at 26-28 weeks was normal, then routine testing of these values at term can be avoided and can result in significant cost savings.^24^ These suggested pre-delivery Hb screening schedules are important as they also provide some opportune time to correct anemia, if detected before labour begins.

Although anemia in pregnancy can have serious consequences for both mother and newborn’s health and well-being throughout their lives, it receives little attention from healthcare practitioners and researchers in low-income nations.^25^ There are no specific policies or guidelines for screening of newborn anemia in Africa. To the best of our knowledge, no other study has addressed this problem locally, and the 6 Public Hospitals involved in this study serve a large number of populations within and around the Central province of the country. Therefore, this study aimed to investigate the timings of antenatal hemoglobin and associated maternal and neonatal characteristics in public hospitals located in Lusaka district of Zambia.

## Materials and methods

The methods employed in carrying out this study are discussed in this section.

### Study Design

This was a secondary analysis of a cross-sectional study titled ‘ASSESSING THE IMPACT OF DELAYED CORD CLAMPING ON NEONATAL HAEMOGLOBIN LEVELS IN VAGINAL DELIVERIES AT THE WOMEN AND NEWBORN HOSPITAL AND PUBLIC FIRST LEVEL HOSPITALS IN LUSAKA DISTRICT, ZAMBIA’ with The Institutional Review Board of the University of Zambia (UNZABREC) approved reference number 4998-2024.

### Study setting

The study was conducted at 6 public hospitals in Lusaka district: the Women and Newborn Hospital and 5 first level hospitals. The Woman and Newborn Hospital is part of the University Teaching Hospitals, which is the largest tertiary hospital in the country and is the highest national referral centre receiving patients from all the 24 local clinics in the Lusaka district (including those from private health facilities). This hospital conducts at least 15,000 deliveries annually. It has a Neonatology Unit with a capacity to admit 60 newborns at once, but it often admits between 90 to 100 newborns, both inborn and out born. Additionally, the Kangaroo Mother Care unit can accommodate 26 neonates. Women and Newborn Hospital offers a range of obstetric and neonatal care, including labour and delivery services, family planning, postnatal care, neonatal intensive care, and support for premature or ill newborns.

The First Level Hospitals included Chawama, Chilenje, Chipata, Kanyama, and Matero hospitals. In the Zambian context, First Level Hospitals are akin to district hospitals and serve as referral centres within their respective constituencies. They are strategically located in the peri-urban areas of Lusaka, catering to the healthcare needs of the local population within their localities. In addition to the general medical and surgical services, the hospitals provide essential maternal and neonatal services, including maternity care and deliveries. Expectant mothers can access skilled birth attendants and receive necessary care during childbirth.^26^

### Sampling Method

Six Research Assistants were recruited in each of the Public Hospitals. Sampling technique was consecutive, on first-come-first-serve basis until the desired sample size of 572 was obtained. Recruitment of participants in all the 6 Public Hospitals took place concurrently. The mother-newborn pair were recruited from the admission wards of the 6 public hospitals located in Lusaka District. The study was conducted over a 5-month period between 15th January to 3rd June 2024 at the 6 public hospitals as outlined in Table 1. Eighty-three (14.5%) out of the 572 participants recruited, had incomplete hospital records, hence only data from 489 mother-newborn pairs were analysed.

**Table 1.** Sample Distribution.

| Setting | Percentage | Sample size |
| --- | --- | --- |
| Women and Newborn Hospital | 40% | 229 |
| First level hospitals | 60% | 343 |
| --Chawama | 12.1% | 69 |
| --Chilenje | 12% | 68 |
| --Chipata | 12.1% | 69 |
| --Kanyama | 12.1% | 69 |
| --Matero | 12% | 68 |
| Total | 100% | 572 |

### Data source and sample

The mother-newborn’s socio-demographic and clinical data were collected using structured questionnaire based-interviews that were created in English and translated to the local languages. Maternal body mass index (BMI) was calculated from measured weight (kg) and height (cm) using the standard method during the interview. Latest maternal Hb and date were obtained from the hospital file after delivery. The weight of the newborn baby was determined by midwives at birth. Timing of umbilical cord clamping after birth was recorded.^27^ Newborn Hemoglobin was determined with a portable hemoglobinometer, the HemoCue Hgb analyzer (HemoCueHb 201+, Sweden) according to standard guidelines within the first 24 hrs after birth. In this study, newborn anemia was defined as Hgb <15g/dL and maternal anemia as Hgb <11g/dL.^28,29^

### Statistical analysis

The data was manually cleaned, processed, checked for completeness and entered into Microsoft Excel. It was then exported into SPSS version 26 for analysis. After categorizing and defining the variables, a descriptive analysis was carried out for each of the independent variables and presented with numbers, frequencies and percentages. Chi test and Binary logistic regression analysis were used to assess the relationship between the timing of antenatal hemoglobin and each independent variable. Multicollinearity and fitness of the model were checked. Factors with p-values <0.1 were included in the regression model. Odds ratio (OR), with a 95% confidence interval (CI) were computed. For all, statistical significance was declared at p-value <0.05. The reporting in this study were guided by the STROBE guidelines for observational studies.^30^

### Ethics

The Institutional Review Board of the University of Zambia (UNZABREC) approved the original study titled ‘ASSESSING THE IMPACT OF DELAYED CORD CLAMPING ON NEONATAL HAEMOGLOBIN LEVELS IN VAGINAL DELIVERIES AT THE WOMEN AND NEWBORN HOSPITAL AND PUBLIC FIRST LEVEL HOSPITALS IN LUSAKA DISTRICT, ZAMBIA’ with reference number 4998-2024, and this was the source of the data for this article. Additional approvals from NHRA, WNH, and Lusaka Provincial Health Office were obtained before data could be collected. Further permission was obtained from Senior Medical Superintendents at each of the first level hospitals. The mother’s written informed consent and assent were obtained. Participants were assured of confidentiality as well as anonymity. The participants were informed that they were free to withdraw from the study without any negative consequences. All study documents were secured under a locked cabinet. This study posed no harm to the participants as it was an observational study. For mothers and newborns diagnosed with anemia, communication was established with the attending Doctor for further assessment and treatment. In order to protect the privacy and confidentiality of the participants, no personal identification such as name was collected. This study was carried out in accordance with the Helsinki Declaration. Results of the study will be made available to stakeholders to obtain information, which could be used for strategies to improve perinatal care.

### Results

The following are the results obtained from the study.

### Participants

The study targeted pregnant women giving birth at WNH-UTH and all the 5 first level Hospitals in Lusaka district.

### Clinical characteristics of the newborns in the study

Of note, only 135 (27.6%) newborns out of the 489 recruited had normal hemoglobin levels of ≥15g/dl and 12 (2.5%) were severely anemic with Hb <10g/dl, Table 2.

**Table 2:**
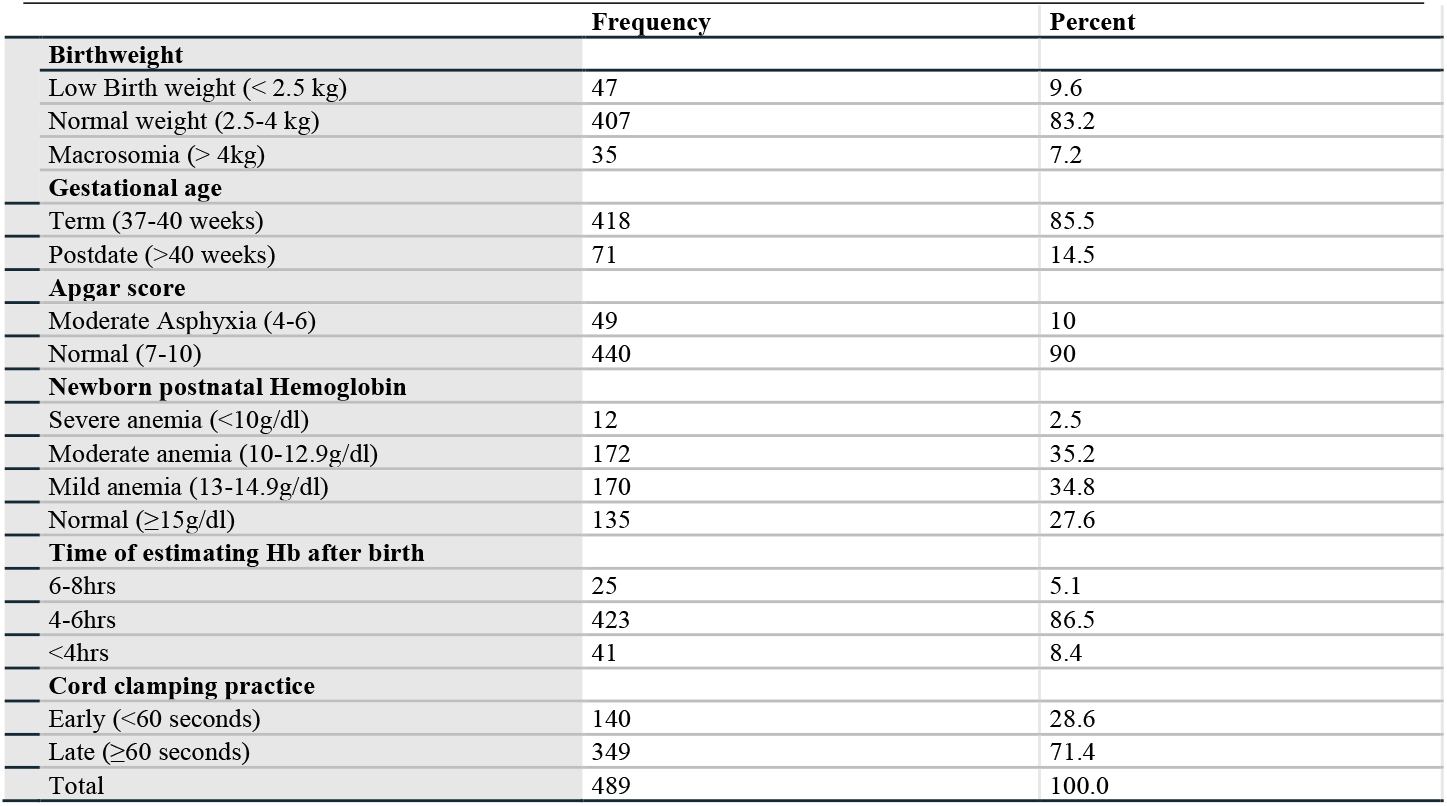
Clinical characteristics of newborns in the study, n=489.

### Characteristics of the mothers in the study

A higher percentage of the mothers were young (>25-35years of age, 43.1%), overweight (47.9%), of secondary school education (50.3%), primiparous (40.7%) with no underlying medical condition (89.0%) and have never had a miscarriage (81.0%). Notably, the adolescents exhibited a higher prevalence (9.4%) of mothers with a low Body Mass Index (BMI), compared to other age groups, p<0.001 (OR 3.84; 95%CI 1.03,14.36), Table 3.

**Table 3:** Maternal characteristics in the study, n=489.

| Maternal Characteristics | Frequency | Percent |
| --- | --- | --- |
| **Age** |  |  |
| Teenage (< 18 years) | 32 | 6.5 |
| Very young (18-25 years) | 209 | 42.7 |
| Young (>25-35 years) | 211 | 43.1 |
| Elderly (>35 years) | 37 | 7.6 |
| **BMI (kg/m<sup>2</sup>)** |  |  |
| Wasted (<18.5) | 15 | 3.1 |
| Normal (18.5-24.9) | 174 | 35.6 |
| Overweight (25-29.9) | 234 | 47.9 |
| Obese (>30) | 66 | 13.5 |
| **Education** |  |  |
| None | 31 | 6.3 |
| Primary | 144 | 29.4 |
| Secondary | 246 | 50.3 |
| Tertiary | 68 | 13.9 |
| **Maternal Medical (present or absent)** |  |  |
| Presence of underlying medical condition | 54 | 11.0 |
| Absence of underlying medical condition | 435 | 89.0 |
| **Parity (delivery)** |  |  |
| 1 | 199 | 40.7 |
| 2 | 108 | 22.1 |
| 3 | 100 | 20.4 |
| 4 | 64 | 13.1 |
| >4 | 18 | 3.7 |
| **Miscarraige (Yes/No)** |  |  |
| Yes | 93 | 19.0 |
| No | 396 | 81.0 |
| Total | 489 | 100.0 |

### Maternal medical conditions in the study

A higher proportion of mothers with underlying medical conditions had HIV in the study (19/54=35.2%), followed by hypertension (14/54=25.9%) and Hepatitis B virus infection (13.0%), Table 4.

**Table 4:** Maternal medical diagnosis in the study, n=54.

| Maternal Medical Diagnosis | Frequency | Percent |
| --- | --- | --- |
| Diabetes Mellitus | 3 | 5.6 |
| Hepatitis B Virus infection | 7 | 13 |
| Heart Disease | 3 | 5.6 |
| HIV infection | 19 | 35.2 |
| Hypertension | 14 | 25.9 |
| Pregnancy Induced Hypertension | 4 | 7.4 |
| Sickle Cell Disease | 4 | 7.4 |
| Total | 54 | 100.0 |

### Timing of antenatal maternal hemoglobin testing and neonatal, maternal characteristics

A large number of mothers (250/489, 51.1%) did not have record of their last antenatal Hb in their files; 156/489 (31.9%) had antenatal Hb documented more than 4 weeks prior to delivery and only 83/489 (17.0%) had documented Hb within 4weeks of delivery. Hence, out of these 489 mothers in the study, only 239 (48.9%) had file record of antenatal hemoglobin. Mothers, with last documented antenatal hemoglobin test of more than 4weeks (156/239; 65.3%) were 2.22 times (OR 95%CI 1.21, 4.07) more likely to be anemic (Hb<11g/dl).

Neonatal hemoglobin and Apgar scores; maternal education, BMI, medical status and history of miscarriage were significantly different amongst the 3 categories of mothers in Table 5. Of note a higher percentage of LBW babies (44.7%) were observed in the category of mothers who had no record of antenatal hemoglobin, compared to those who had.

**Table 5:**
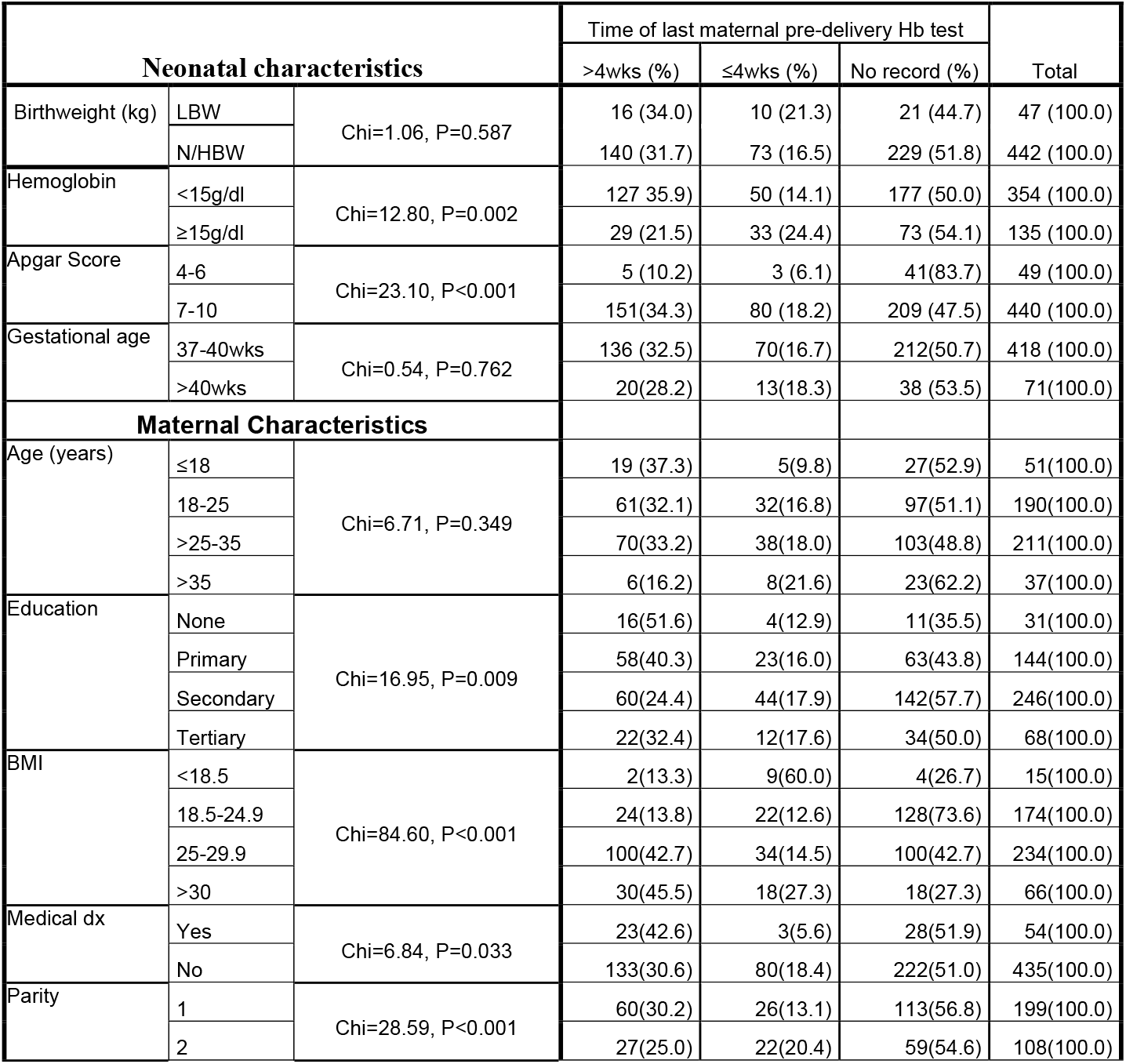
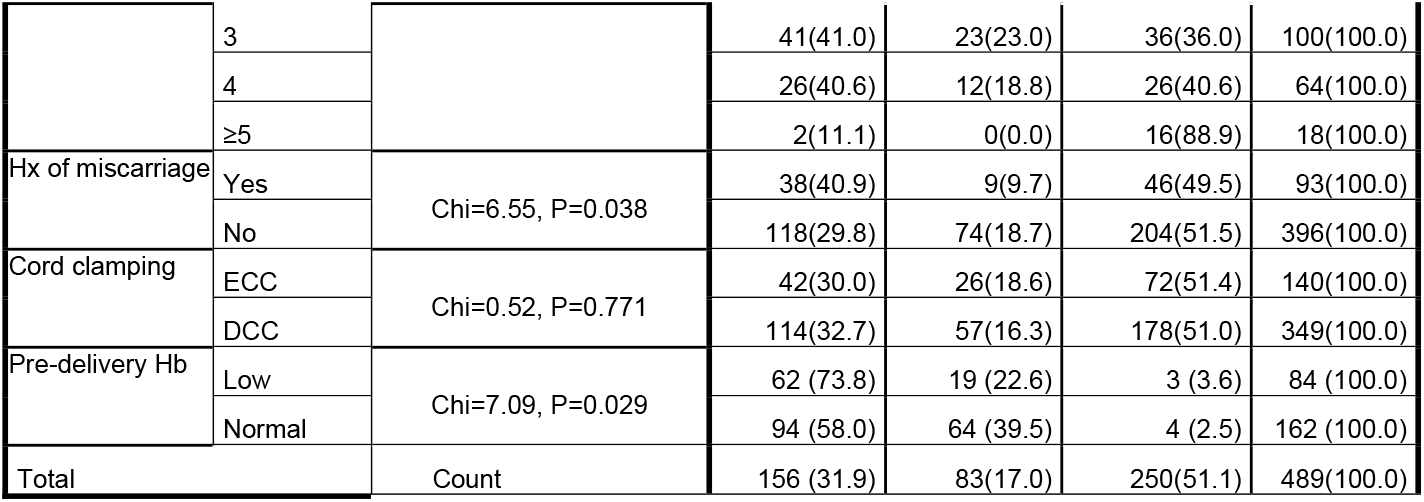
Timing of antenatal hemoglobin test and neonatal, maternal characteristics.

### Binary logistic regression analysis of neonatal and maternal characteristics associated with absence or presence of file record of antenatal hemoglobin test

Table 6 shows that maternal BMI was the strongest and only significant factor associated with the absence or presence of documented antenatal hemoglobin test (Wald 28.907), after conducting a binary logistic regression analysis. Tables 5 and 6 show that lack of documented antenatal hemoglobin test was less likely in mothers with overweight/obese BMI. Mothers with lower BMI were 2.14 times (95% CI:1.62, 2.82) more likely to lack antenatal hemoglobin testing, compared to those with higher BMI in this study, p<0.001.

**Table 6:**
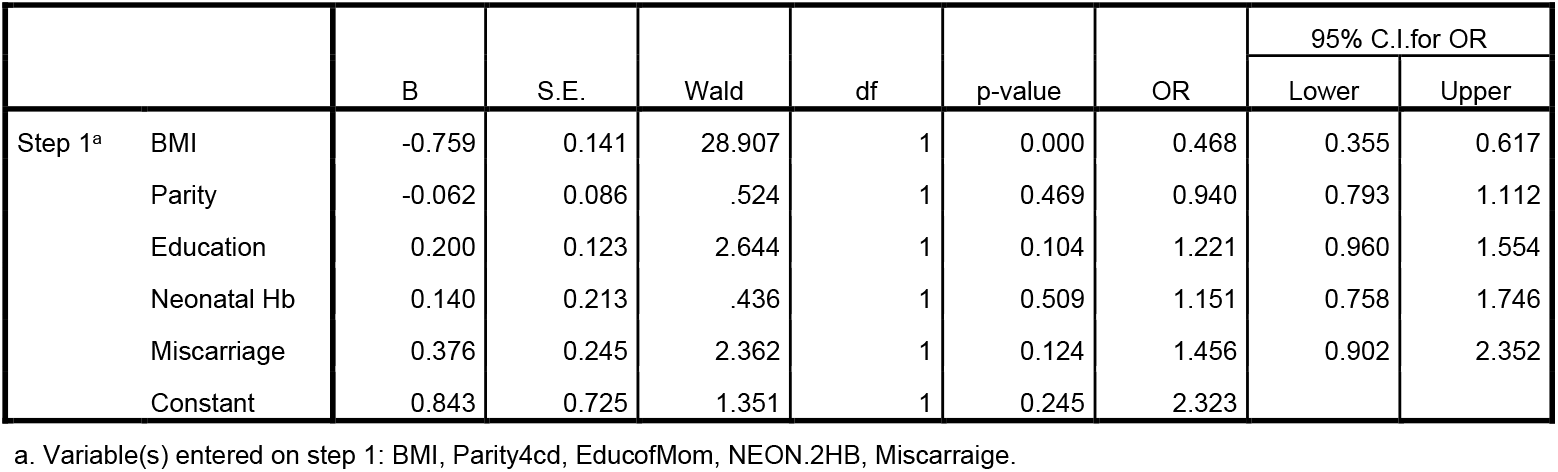
Binary logistic regression analysis of neonatal and maternal characteristics associated with absence or presence of file record of antenatal hemoglobin test result, n=489.

### Binary logistic regression analysis of neonatal and maternal characteristics associated with file record of antenatal hemoglobin within or beyond 4 weeks prior to delivery

#### Maternal BMI

Table 7 shows that maternal BMI was again the strongest factor determining the timing of antenatal hemoglobin testing (Wald 22.728) during gestation. The presence of file record of hemoglobin test result within 4 weeks prior to delivery was less likely in mothers with the higher maternal BMI (OR 0.291; 95% CI 0.175, 0.484). Mothers with lower BMI were 3.44 times (95% CI 2.07, 5.71) more likely to have their antenatal hemoglobin testing done within 4 weeks prior to delivery, compared to those with higher BMI in this study, p<0.001. In Table 5, the higher the maternal BMI, the more likely the last hemoglobin test was done beyond 4weeks prior to delivery. Of particular note, the overweight mothers were 13.16 times (95% CI 2.72, 62.5) more likely to test beyond 4weeks prior to delivery, compared to others. In Table 5, 30 out of 66 (45.5%) mothers with obese BMI had their last hemoglobin test done beyond 4weeks prior to delivery.

**Table 7:** Binary logistic regression analysis of neonatal and maternal characteristics associated with antenatal hemoglobin testing within or beyond 4 weeks prior to delivery, n=239.

|  | B | S.E. | Wald | df | Sig. | Exp(B) | 95% C.I. for EXP(B) |  |
| --- | --- | --- | --- | --- | --- | --- | --- | --- |
|  |  |  |  |  |  |  | Lower | Upper |
| Step 1<sup>a</sup> BMI | -1.233 | 0.259 | 22.728 | 1 | 0.000 | 0.291 | 0.175 | 0.484 |
| Parity | 0.107 | 0.146 | .538 | 1 | 0.463 | 1.113 | 0.836 | 1.483 |
| Apgar score | 1.245 | 0.809 | 2.371 | 1 | 0.124 | 3.475 | 0.712 | 16.957 |
| Education | 0.483 | 0.199 | 5.880 | 1 | 0.015 | 1.622 | 1.097 | 2.397 |
| Neonatal hemoglobin | 1.358 | 0.389 | 12.174 | 1 | 0.000 | 3.889 | 1.813 | 8.338 |
| Maternal medical conditn | 1.641 | 0.726 | 5.100 | 1 | 0.024 | 5.158 | 1.242 | 21.423 |
| Hx of Miscarriage | 1.996 | 0.538 | 13.772 | 1 | 0.000 | 7.362 | 2.565 | 21.131 |
| Antenatal maternal Hb | 1.395 | 0.392 | 12.680 | 1 | 0.000 | 4.037 | 1.873 | 8.701 |
| Constant | -13.251 | 3.457 | 14.696 | 1 | 0.000 | 0.000 |  |  |

#### History of miscarriage/s

Table 7 shows that history of past miscarriage/s was the next strongest factor determining the timing of antenatal hemoglobin testing (Wald 13.772; p<0.001; OR 7.362; 95%CI 2.565, 21.131). Those who had positive history of previous miscarriage were less likely to have their hemoglobin test done within 4 weeks prior to delivery OR 0.378 (p=0.015; 95% CI 0.173, 0.826). In Table 5, 38 out of 93 (40.8%) mothers with positive history of previous miscarraige/s had their last hemoglobin test done beyond 4weeks prior to delivery.

#### Number of miscarriages and file record of antenatal hemoglobin

In Table 8, a higher percentage of mothers who had a positive history of miscarriage and those who have had multiple previous miscarriages were in the group that did not have any file record of antenatal hemoglobin. The mothers who have had >1 previous miscarriages (18 out of 26, 69.2%, p=0.018) were 3.134 times (95%CI 1.195, 8.218) more likely to be in the category of mothers with no file record of antenatal hemoglobin test.

**Table 8:**
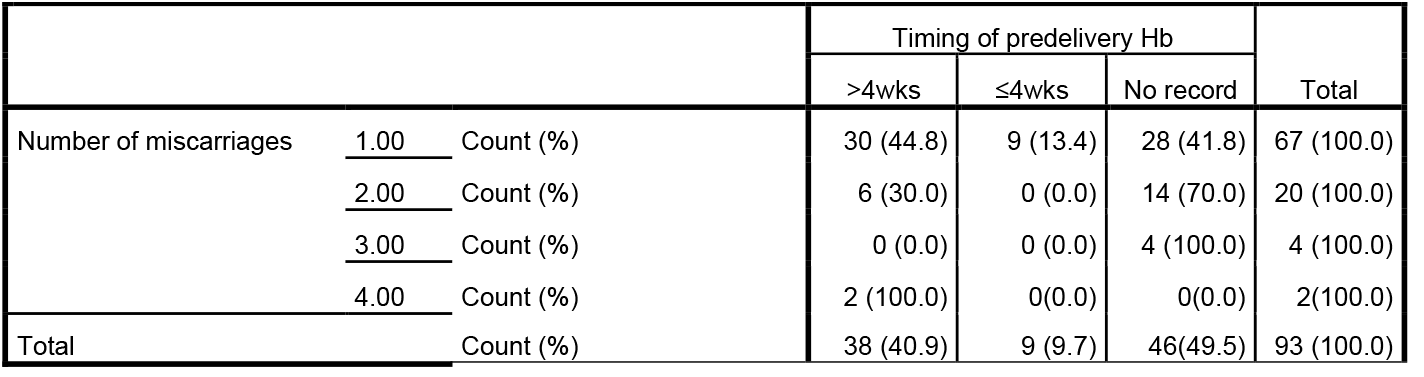
Crosstabulation of number of miscarriages and file record of antenatal hemoglobin, n=93.

|  |  |  | Timing of predelivery Hb |  |  | Total |
| --- | --- | --- | --- | --- | --- | --- |
|  |  |  | >4wks | ≤4wks | No record |  |
| Number of miscarriages | 1.00 | Count (%) | 30 (44.8) | 9 (13.4) | 28 (41.8) | 67 (100.0) |
|  | 2.00 | Count (%) | 6 (30.0) | 0 (0.0) | 14 (70.0) | 20 (100.0) |
|  | 3.00 | Count (%) | 0 (0.0) | 0 (0.0) | 4 (100.0) | 4 (100.0) |
|  | 4.00 | Count (%) | 2 (100.0) | 0(0.0) | 0(0.0) | 2(100.0) |
| Total |  | Count (%) | 38 (40.9) | 9 (9.7) | 46(49.5) | 93 (100.0) |

#### Maternal antenatal hemoglobin level and file record

Level of maternal antenatal hemoglobin was significantly associated with the timing of hemoglobin testing (wald 12.680; p<0.001; OR 4.037; 95%CI 1.873, 8.701) in Table 7. Mothers with low antenatal hemoglobin (Hb<11g/dl) were less likely to be tested within 4weeks prior to delivery (p=0.010; OR =0.45; 95%CI 0.246, 0.824) compared to those with normal hemoglobin. In Table 5, 62 out of 84 (73.8%) mothers with low hemoglobin were last tested more than 4weeks prior to delivery, compared to 94 out of 162 (58.0%) of those with normal hemoglobin.

#### Neonatal hemoglobin level and file record of maternal antenatal hemoglobin

Level of neonatal hemoglobin was also significantly associated with timing of maternal antenatal hemoglobin testing (wald 12.174; p<0.001; OR 3.889; 95%CI 1.813, 8.338) in Table 7. The higher the level of neonatal hemoglobin at birth, the more likely maternal antenatal hemoglobin was tested within 4 weeks. In Table 5, 127 out of 354 (35.9%) babies with low hemoglobin levels (<15g/dl) belonged to mothers, whose last antenatal hemoglobin was tested more than 4weeks prior to delivery (p<0.001; OR 2.89; 95%CI 1.590, 5.236), compared to 29 out of 135 (21.5%) of babies with normal hemoglobin levels.

#### Maternal education and file record of antenatal hemoglobin

Level of maternal education was significantly associated with timing of maternal antenatal hemoglobin testing (Wald 5.880; p=0.015; OR 1.622; 95%CI 1.097, 2.397) in Table 7. The lower the educational level of the mother, the more likely the last antenatal hemoglobin was tested beyond 4weeks (OR 2.182; OR 0.593, 8.022). In Table 5, 16 out of 31 (51.6%) of mothers with no formal education had last antenatal hemoglobin tested beyond 4weeks, compared to 22 out of 68 (32.4%) of mothers with tertiary education.

#### Maternal medical status and file record of antenatal hemoglobin

Maternal medical status was significantly associated with timing of antenatal hemoglobin testing (Wald 5.100; p=0.024; OR 5.158; 95%CI 1.242, 21.423) in Table 7. Those with underlying medical conditions were 4.612 times (95%CI 1.342, 15.851; 23 out of 26; 88.5%) more likely to have their last antenatal hemoglobin tested beyond 4weeks, compared with those who had no underlying medical condition (133 out of 213, 62.4%), Table 5. In Table 9, maternal HIV was the highest medical condition (35.2%) followed by hypertension (25.9%). Six out of these 7 documented types of medical conditions either had remote antenatal hemoglobin or did not have any file record of hemoglobin. Only diabetes mellitus was within 4 weeks of predelivery hemoglobin testing.

**Table 9:**
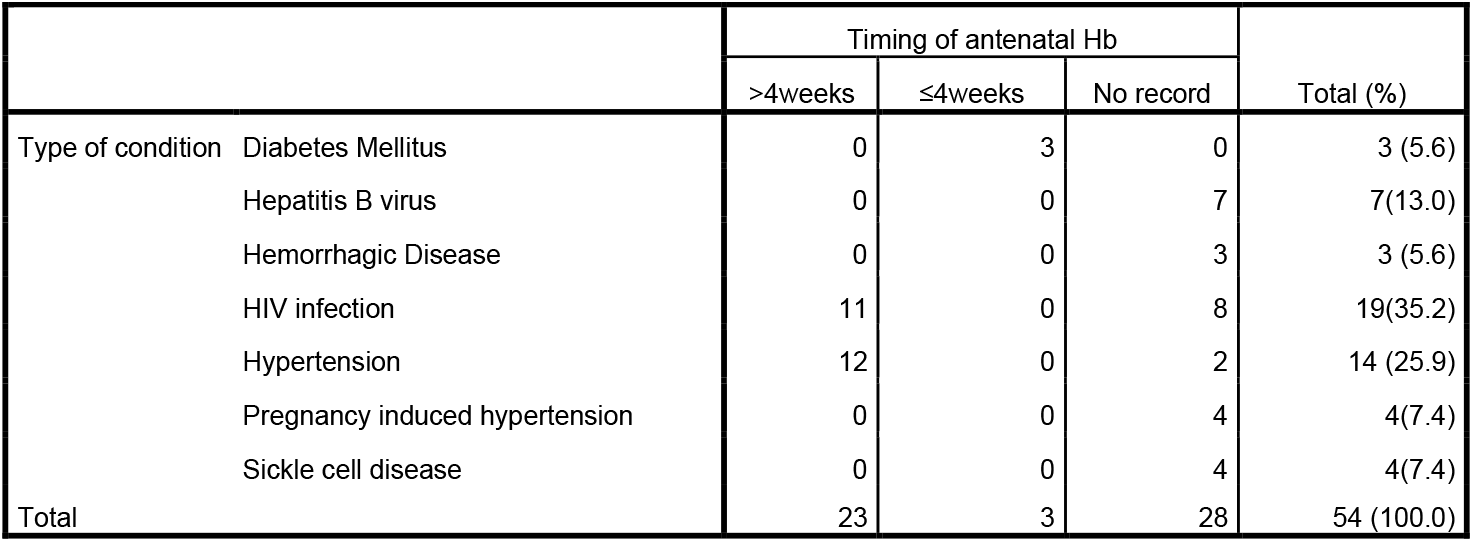
Crosstabulation of maternal medical condition and file record of antenatal hemoglobin, n=54.

|  |  | Timing of antenatal Hb |  |  | Total (%) |
| --- | --- | --- | --- | --- | --- |
|  |  | >4weeks | ≤4weeks | No record |  |
| Type of condition | Diabetes Mellitus | 0 | 3 | 0 | 3 (5.6) |
|  | Hepatitis B virus | 0 | 0 | 7 | 7(13.0) |
|  | Hemorrhagic Disease | 0 | 0 | 3 | 3 (5.6) |
|  | HIV infection | 11 | 0 | 8 | 19(35.2) |
|  | Hypertension | 12 | 0 | 2 | 14 (25.9) |
|  | Pregnancy induced hypertension | 0 | 0 | 4 | 4(7.4) |
|  | Sickle cell disease | 0 | 0 | 4 | 4(7.4) |
| Total |  | 23 | 3 | 28 | 54 (100.0) |

#### Miscellaneous

Maternal age, parity, cord clamping, neonatal apgar score and gestational age were not significantly associated with timing of hemoglobin testing. Teenage mothers (<18yrs) [p=0.028; OR 5.08; 95%CI 1.19, 2.74] and young mothers (>25-35years) [p=0.048; OR 2.60; 95%CI 1.0, 6.71] were more likely to have their antenatal hemoglobin performed beyond 4weeks.

Newborns with low apgar score 4-6 (moderate asphyxia) were more likely to be associated with mothers with no record of antenatal hemoglobin (p<0.001; OR 5.924; (95% CI 2.287, 15.345) compared to those with normal apgar score.

Para 3 mothers were more likely [p=0.109; OR 1.709; 95%CI 0.887, 3.289] to have their antenatal hemoglobin performed beyond 4weeks.

The early term (gestation 37-40weeks) babies were more likely to be associated with mothers with remotely timed antenatal hemoglobin testing [p=0.545; OR 1.260; 95%CI 0.593, 2.90].

Early cord clamping was associated with antenatal hemoglobin performed within 4weeks of delivery [p=0.473; OR 1.238; 95%CI 0.691, 2.219].

Low birthweight babies were more likely to be associated with mothers who had their predelivery Hb test conducted within 4 weeks prior to delivery [p=0.672; OR 1.199; 95%CI 0.518, 2.774].

## Discussion

The study investigated the characteristics of mothers and their newborns within a sample of 489 term, vaginal deliveries, with particular emphasis on the timing of the last antenatal hemoglobin testing. The results indicated that a noteworthy 51.1% of mothers did not have documentation of hemoglobin tests in their hospital records at the time of delivery. Additionally, 31.9% of mothers had their most recent antenatal hemoglobin testing conducted more than four weeks prior to delivery.

Mothers who either were not tested or tested infrequently for anemia during pregnancy, exhibited several concerning characteristics, as expected. These include low educational attainment, low body mass index (BMI), a history of previous miscarriages, low hemoglobin levels, and underlying medical conditions, in addition to newborns presenting with low hemoglobin levels. Of particular significance is the observation that mothers with a history of multiple miscarriages demonstrated a marked tendency to lack documentation of antenatal hemoglobin testing.

Maternal body mass index (BMI) has been identified as a significant factor influencing pregnant women’s engagement with antenatal care. This conclusion is based on a retrospective analysis of routine hospital data from 34 NHS maternity units in England, United Kingdom, which examined 619,502 singleton births between 1989 and 2007. The findings indicated that mothers who were classified as overweight or obese tended to access antenatal care later in their pregnancies compared to those with lower BMIs. Furthermore, it was observed that mothers from minority ethnic groups, teenagers, the unemployed, and those experiencing deprivation, particularly Black and Black British women, often sought care only during the third trimester.^31^

Conversely, this current study conducted in Zambia revealed that mothers with low BMI were more likely to lack documented antenatal hemoglobin testing. However, when there is a documented record of hemoglobin testing, it appears that mothers with lower body mass indexes (BMIs) tend to undergo testing at earlier stages of gestation. Conversely, mothers classified as overweight or obese tend to undergo testing later in their pregnancy, which aligns with the findings noted in the study conducted in the United Kingdom.

These findings suggests a potential deficiency in engagement with antenatal care services, generally. Additionally, a secondary analysis of the 2018 Zambia Demographic Health Survey, conducted by Ahmed, indicated that only approximately 64.1% of pregnant mothers utilized antenatal services, despite the fact that these services are offered at no cost in Zambia. Notably, low socioeconomic status and rural residency emerged as significant factors constraining adequate use of these vital antenatal services.^32^

The highest prevalence of anemia during pregnancy occurs in Sub-Saharan Africa and South Asia, with rates of 41.7% and 40%, respectively. A secondary analysis of the 2015 to 2022 Demographic Health Survey (DHS) dataset, encompassing 14,098 pregnant women across 21 Sub-Saharan African countries, identified Mali as having the highest prevalence of anemia in pregnancy at 69.21%, followed by Côte d’Ivoire at 68.21%.^33^ In comparison, a previous paper reporting on anemia in pregnancy from these same six public hospitals in Lusaka, Zambia, presented a prevalence of 30.5%,^34^ which is significantly lower than the figures observed in Mali and Côte d’Ivoire.

Despite these high levels of anemia in pregnancy, particularly in this sub-Saharan Africa region, hemoglobin measurement during pregnancy does not appear to be routine in several health facilities in this study and likewise in a mixed-methods study of situational analysis of facility readiness and community understanding of anemia in Ghana and Uganda.^35^ Even though it is widely acknowledged that timely antenatal hemoglobin testing is essential for the prompt identification and correction of anemia prior to delivery. This is particularly critical for women who may have experienced irregular attendance at antenatal clinics and do not possess a recent hemoglobin measurement.

Antenatal maternal hemoglobin testing constitutes one of the six essential examinations recommended by the World Health Organization (WHO) as part of standard care for pregnant women. The other tests include screening for proteinuria, syphilis, sickle cell anemia and HIV, blood group determination.^36^ Implementing these tests is crucial for promoting safe delivery and ensuring the health and well-being of both the mother and the newborn.

The findings in this study suggest possible deficiencies in the quality of antenatal care provided by the public health facilities assessed in this study. The equipment and materials necessary for a comprehensive antenatal screening panel may be available in the laboratories of hospital-level facilities within the public health care system. However, numerous lower-level health facilities, which are more accessible to communities and focus on outpatient services and uncomplicated deliveries, frequently lack the laboratory resources required to conduct the full spectrum of laboratory tests needed during pregnancy.^37^ A quality assessment study on antenatal care in Zambia, conducted by Kyei et al. in 2012 using data from the 2005 Zambia Health Facility Census and the 2007 Zambia Demographic and Health Survey, involved 1,299 antenatal facilities and 4,148 mothers. The findings revealed that merely 3% of health facilities provided optimal antenatal care services, while 47% offered adequate care, and 50% rendered inadequate services. Although 94% of mothers reported at least one visit with a qualified health worker, only 29% received good quality antenatal care, and just 8% received high-quality care after beginning their visits in the first trimester.^38^

Various other factors, such as mother’s economic status, distance from the nearest health facility, late presentation for antenatal care, limited point-of-care testing options, inadequate staffing levels, and the lack of standardized guidelines for the management of anemia in pregnancy, have also contributed to insufficient provision of antenatal testing. Consequently, not all pregnant women receive hemoglobin testing at each antenatal care visit as a standard practice.^35^

This study highlights the critical importance of delivering high-quality antenatal care, inclusive of routine hemoglobin testing, to facilitate the timely identification and management of anemia, comorbidities and complications. Such measures are essential for promoting positive maternal experiences, ensuring safe deliveries, and fostering the health of newborns.

### Limitation

The available data lacked specific details concerning antenatal care.

#### Strength

This study, conducted across multiple centers, yielded valuable insights into the antenatal care practices, maternal and newborn characteristics in resource-limited settings, addressing a gap in the current literature.

#### Conclusion ad Recommendations

The campaign for comprehensive antenatal care, particularly for pregnant mothers in resource-limited settings, should be significantly enhanced. The objective is to prevent maternal and newborn anemia, thereby contributing to the reduction of maternal and infant morbidity and mortality rates.

## Data Availability

All data produced in the present study are available upon reasonable request to the authors

## Author Contributions

The corresponding author (Dr Adenike Oluwakemi Ogah) co-supervised the study and the original dissertation, conducted the secondary data analysis, interpreted the results and drafted this manuscript. Dr Chrispin Mwando, conceived the study title, collected the data and drafted the original dissertation. Dr Kenneth Chanda supervised the study and original dissertation. Dr Selia Nganjo co-supervised the study and original dissertation. All the authors contributed to the intellectual content and final editing of this manuscript.

## Acknowledgements

The authors are extremely grateful to the participants involved in this study, to the staff of each of the Public Hospitals in Lusaka that contributed to this study and to the research team.

